# Interictal SEEG resting-state connectivity localizes seizure onset zone and predicts seizure outcome

**DOI:** 10.1101/2021.12.30.21268524

**Authors:** Haiteng Jiang, Vasileios Kokkinos, Shuai Ye, Alexandra Urban, Anto Bagić, Mark Richardson, Bin He

**Author notes:** Correspondence: Bin He, PhD, Department of Biomedical Engineering, Carnegie Mellon University, 5000 Forbes Avenue, Pittsburgh, PA 15213.

## Abstract

Stereotactic-electroencephalography (SEEG) is a common neurosurgical method to localize epileptogenic zone in drug resistant epilepsy patients and inform treatment recommendations. In the current clinical practice, localization of epileptogenic zone typically requires prolonged recordings to capture seizure, which may take days to weeks. Although epilepsy surgery has been proven to be effective in general, the percentage of unsatisfactory seizure outcomes is still concerning. We developed a method to identify the seizure onset zone (SOZ) and predict seizure outcome using short-time resting-state SEEG data. In a cohort of 43 drug resistant epilepsy patients, we estimated the information flow via directional connectivity and inferred the excitation-inhibition ratio from the 1/f power slope. We hypothesized that the antagonism of information flow at multiple frequencies between SOZ and non-SOZ underlying the relatively stable epilepsy resting state could be related to the disrupted excitation-inhibition balance. We found higher excitability in non-SOZ regions compared to the SOZ, with dominant information flow from non-SOZ to SOZ regions, probably reflecting inhibitory input from non-SOZ to prevent seizure initiation. Greater differences in information flow between SOZ and non-SOZ regions were associated with favorable seizure outcome. By integrating a balanced random forest model with resting-state connectivity, our method localized the SOZ with an accuracy of 85% and predicted the seizure outcome with an accuracy of 77% using clinically determined SOZ. Overall, our study suggests that brief resting-state SEEG data can significantly facilitate the identification of SOZ and may eventually predict seizure outcomes without requiring long-term ictal recordings.

## Introduction

Epilepsy is one of the most common neurological diseases (*1*) impacting about 70 millions people in the world. At least one-third of epilepsy patients becoming drug-resistant and potential candidates for surgical resection or neuromodulation (*2*). The key to successful epilepsy surgery relies on accurate localization and safe removal of the epileptogenic zone (EZ) (*3*) and an understanding of an individual patient’s seizure network (*4, 5*). An integral component for the delineation of the EZ is the seizure onset zone (SOZ): the area of cortex that initiates clinical seizures as determined predominantly by intracranial investigations (*6, 7*). Although surgery and neuromodulation have been proven efficient in seizure reduction, the percentage of patients with unfavorable seizure outcomes leaves significant room for improvement (*8*).

SEEG is a well-established and safe neurosurgical approach (*9*) to identify epileptic regions for intervention with intracerebral electrodes to record ictal/interictal brain activity (*10-13*). The golden standard of localization of epileptogenic brain regions in clinical practice typically depends on capturing multiple seizures during the intracranial monitoring process, that may take multiple days or even weeks to complete (*14*). As such, a method which can estimate SOZ and predict prognosis outcome from analysis of brief, resting-state data segments would have tremendous clinical values to identify epileptogenic networks without requiring prolonged intracranial recordings, which would vastly improve patient care and reduce medical cost (*15-17*).

In a healthy state, the balanced excitation and inhibition in brain networks is regulated to facilitate information transfer and communications between remote functional regions (*18*). A number of studies have indicated that neuronal oscillations could transfer information at different frequencies, and oscillatory dysfunction has been implicated in almost every major psychiatric and neurological disorders (*19, 20*). More specifically, it has been demonstrated that low-frequency activity (LFA, <30 Hz), high-frequency activity (HFA, >30 Hz), and LFA to HFA cross-frequency interactions of the epilepsy network are disrupted (*21-23*). For example, high-frequency oscillations (HFOs) (*24*), interictal epileptiform discharges (IEDs) (*25*), and phase-amplitude coupling (PAC) (*26*) have been widely investigated as promising clinical biomarkers for epilepsy (*27, 28*). However, HFOs, IEDs and PAC are all local biomarkers, while epilepsy is commonly accepted as a network disease (*29*). The underpinnings of seizure generation involve abnormal brain structures and aberrant functional connections among these regions, leading to large-scale network instability (*30, 31*). Resting-state network connectivity studies have suggested predominantly increased functional connectivity involving the EZ and surrounding structures (*15*), and stronger inward directional connectivity toward EZ (*16*). Furthermore, decreased interictal network synchrony and local heterogeneity was found to correlate with improved seizure outcome (*32*). Therefore, a better understanding of the functional architecture of the epileptic network could help identify SOZ and improve prediction of the seizure outcome.

In this work, we investigate information flow in resting-state epilepsy networks, inferred from directional connectivity in a cohort of 43 drug-resistant focal epilepsy patients. We hypothesized that the excitation-inhibition balance is disrupted during epilepsy resting state compared to the healthy resting state and further reflected by aberrant information flow. Specifically, we hypothesized that during the relatively stable epilepsy resting state, there are antagonisms of information flow between SOZ and non-SOZ regions at multiple frequencies. Furthermore, we speculated that the strength of antagonisms reflects intrinsic epileptic network characteristic, which is eventually associated with seizure outcome. The ultimate goal of this work is to develop a method to identify the SOZ for treatment intervention and to predict treatment outcomes, based on brief resting-state SEEG data without necessitating prolonged ictal recordings (Fig. 1).

**Fig. 1.**
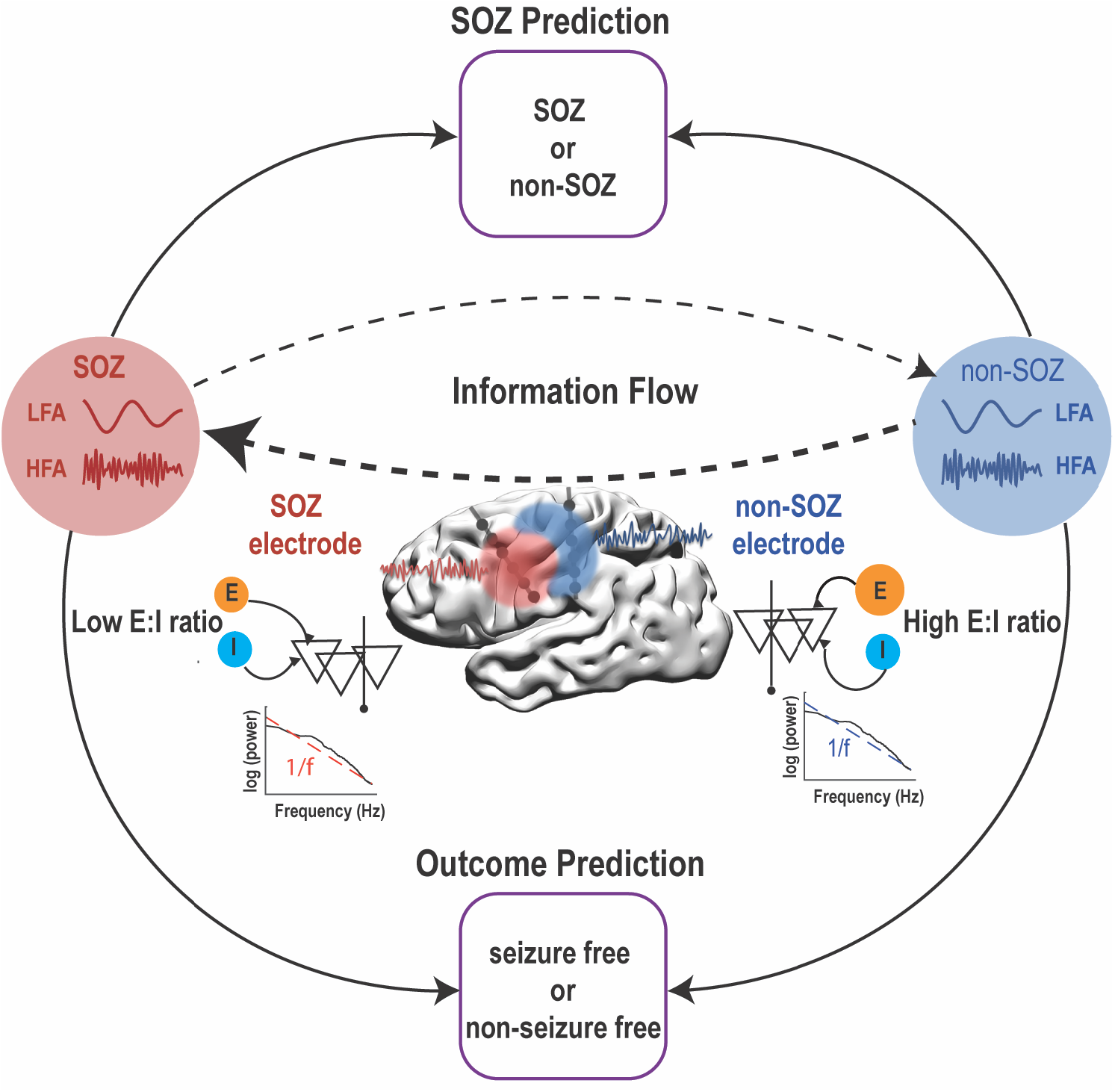
Schematic illustration of the study design. Within-frequency and cross-frequency directional connectivity (indication of information flow), 1/f power slope (indication of exitation and inhibiton ratio) were investigated in the SEEG resting state data to predict SOZ and seizure outcome. LFA: Low-freqeuncy activity; HFA: High-freqeuncy activity; SOZ: Seizure-onset zone; E:I: Excitation:Inhibition.

## Methods and Materials

### Patients

The study included 43 drug-resistant focal epilepsy patients who underwent complete presurgical evaluation, including SEEG at the University of Pittsburgh Medical Center between 2014 and 2019. All patients enrolled during the period with SEEG recordings were considered, and patients with clear recordings of SOZ were included. Demographic and clinical information of patients is summarized in Table S1. Treatments such as surgical resection (19 patients), ablation (8 patients), vagus nerve stimulation (VNS, 2 patients), responsive neurostimulation (RNS, 9 patients), and combined strategies (5 patients) were conducted during the medical care. Besides, postoperative seizure outcome was evaluated at the last follow-up (>1 year) using the Engle classification scale (*33*). This study was approved by local institutional review boards at the University of Pittsburgh and Carnegie Mellon University. Written informed consent was obtained from all patients.

### SEEG data collection

The SEEG data were recorded using the Xltek acquisition system (Natus Medical Inc, Pleasanton, CA) with a 2 kHz or 1 kHz (3 patients) sampling rate. Ten-minute epochs were randomly selected from the long-term SEEG recordings during interictal periods in which the patient was at rest. All selected epochs were recorded between 7:00AM and 12:30PM, and at least 2 hours away from an ictal event. The selected recordings were visually examined for the presence of epileptiform activity. If such activity was observed, the resting epoch was discarded, and another epoch was randomly selected. On average, three such epochs were obtained for each patient. Raw data were notch filtered at 60 Hz and re-referenced using bipolar montage. Electrode pairs residing in white matter were excluded from further analysis.

SOZ was marked by board-certificated epileptologists using established clinical interpretation. The SOZ determination occurred during the intracranial recording session and was completed before any data analysis in this study. The seizure onset was defined by the first electrographic changes, compared to the interictal background patterns, leading to a sustained rhythmic discharge followed by or concurrently related to typical clinical semiology on simultaneous video-EEG (*34, 35*).

### Directional connectivity estimation

The within-frequency and cross-frequency directional information flow were estimated by the directed transfer function (DTF) (*36, 37*) and cross-frequency directionality (CFD) (*38, 39*) respectively. Based on the framework of multivariate autoregressive (MVAR) models, DTF provides a spectral measure for directed information flow in the spectral domain in the multivariate system (*40, 41*). It has been demonstrated that DTF was useful in objectively determining underlying pathological connections such as epilepsy (*42-44*).

Since DTF is only able to estimate the directional information flow at a single frequency, CFD is further utilized to quantify the directional interactions between different frequencies (*39*). CFD has been applied in different electrophysiological modalities, including magnetoencephalography (*46*), electroencephalography (*47, 48*), and electrocorticography (*38, 49*), often revealing new insights into multilayer network interactions. The core basis of CFD is the phase-slope index (PSI), assuming that constant lag in the time lag could be represented by linearly increasing or decreasing phase differences in the considered frequency range (*50*). By computing the PSI between the phase of low-frequency activity (LFA) (< 30 Hz) and the amplitude of high-frequency activity (HFA) (> 30 Hz), the positive CFD indicates information flow from LFA to HFA and vice versa for the negative CFD.

### 1/f power slope estimation

The power-law exponent (slope) of the power spectrum (1/f) has been suggested to estimate synaptic excitation (E)-inhibition (I) ratios, with a flattened power slope indicating high E:I ratio and vice versa for a steep power slope (*51*). Here, we estimated the 1/f power slope with FOOOF package. To obtain power spectrum, data were epoched into 1-second segment without overlapping, and the time-frequency decomposition was estimated by a Fast Fourier Transformation in combination with a Hanning taper from 1 Hz to 250 Hz in 1 Hz step. Then, the power spectrum of each epoch was computed and subsequently averaged over all epochs. After the power spectrum calculation, the FOOOF algorithm operates on power spectrum densities in semilog-power space, which are linearly spaced frequencies and log-spaced power values (*52*). Essentially, the 1/f power slope is fit as a function across the selected range of the spectrum, and each oscillatory peak is modeled with a Gaussian function individually.

### Random forest classification

To predict at the individual electrode level (e.g., SOZ vs. non-SOZ) and patient-level (e.g., seizure-free vs. non-seizure free outcome), we utilized the random forest machine learning technique. Random forest is an ensemble machine learning method that induces each constituent decision tree from bootstrap samples of the training data (*53*). The prediction is made by aggregating all decision trees’ predictions. For SOZ individual prediction, the majority of electrodes are non-SOZ. Thus, these two classes (SOZ and non-SOZ) are severely imbalanced. Dealing with highly imbalanced data, a sample may contain few or even none of the minority class, resulting in a tree with a poor predicting performance for the minority class. To tackle the problem of severely imbalanced data, a balanced random forest was introduced by adopting two strategies: 1) minimizing the overall cost by assigning a high cost to the misclassification of minority class; 2) either over-sampling the minority class or down-sampling the majority class or both (*54*). Here, we applied a synthetic minority over-sampling technique (SMOTE), a combination of over-sampling the minority class and under-sampling the majority class (*55*). The balanced random forest is implemented in open-source python toolbox imbalanced-learn (https://github.com/scikit-learn-contrib/imbalanced-learn) and adapted in our study (*56*). To evaluate the model’s performance, we applied the five-fold cross-validation approach, further generated the receiver operator characteristic (ROC) curve, and computed the area under the curve (AUC). We also assessed the metrics of precision, recall, and overall accuracy.

## Results

### Dominant information flow from non-SOZ to SOZ underlying the resting-state

First, we examined the confrontations of information flows between SOZ and non-SOZ by comparing their differences in directional interaction both within-frequency and cross-frequency. The within-frequency directional information flow was calculated by DTF (*40*), while the cross-frequency directional information was estimated by CFD (*39*). As shown in Fig. 2A, measures of within-frequency information flow strength were significantly weaker from SOZ to non-SOZ than in the other direction, over the wide frequency range (1∼250 Hz). Furthermore, we observed that SOZ exhibited significantly higher inward strength (mean information received from other electrodes) than non-SOZ (Fig. 2B), but outward strength (mean information sent to other electrodes) did not differ between the regions (Fig. 2C). Note that the patterns were similar and robust when the within frequency directional interaction was estimated by partial directed coherence (PDC) (Fig. S1), which is claimed to be more accurate in determining the directed connectivity compared to DTF (*57*). In the cross-frequency directional interactions, both the SOZ phase to non-SOZ amplitude and non-SOZ phase to SOZ amplitude CFD showed prominent negative 1-4 Hz to 40-150 Hz CFD (Fig. 3A), indicating information flow from HFA to LFA. Since CFD varied across electrode pairs to a different extent, we utilized k-means clustering to extract the most consistent and strongest CFD pattern across all electrode pairs (*38*). After applying the k-means procedure in each patient, we found the significant negative CFD from SOZ phase to non-SOZ amplitude CFD (Fig. 3B), suggesting the dominant cross-frequency information flow from non-SOZ HFA to SOZ LFA. Overall, the within-frequency and cross-frequency results indicated that the prevailing information flow is always from non-SOZ to SOZ.

**Fig. 2.**
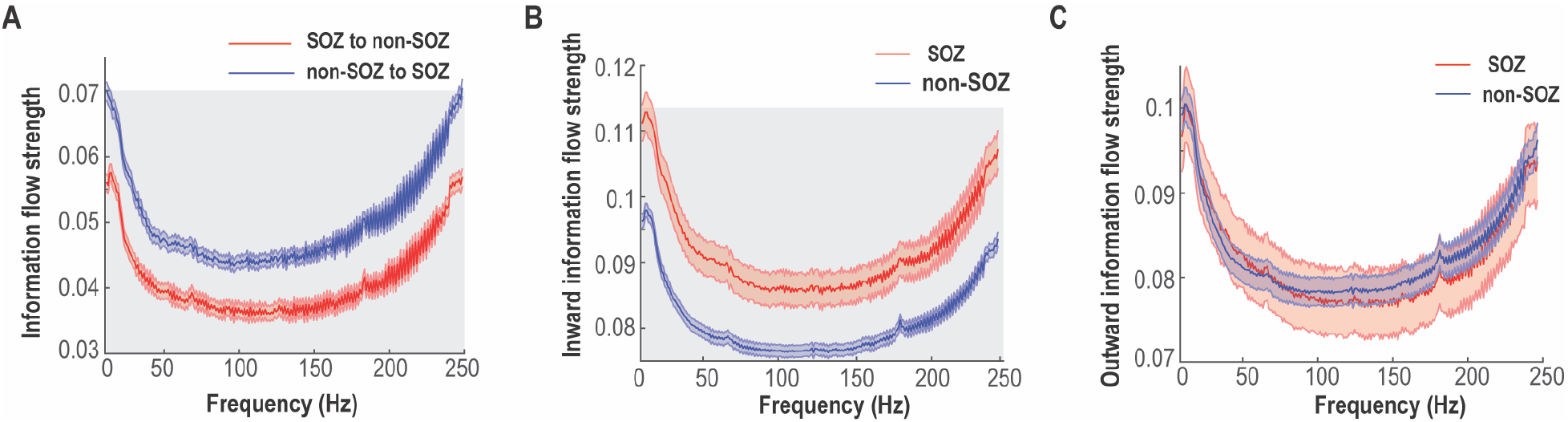
Within-frequency information flow by means of Directed Transfer Function analysis. (A) Mean bidirectional information flows between SOZ and non-SOZ across all electrode pairs and patients. The shaded gray area indicates significant differences at the *p*=0.01 level after multiple corrections. (B) Inward (receiving) information flow strength in SOZ and non-SOZ. The shaded gray area indicates significant differences at the *p*=0.01 level after multiple corrections. (C) Outward (sending) information flow strength in SOZ and non-SOZ. Data are shown in mean and standard error.

**Fig. 3.**
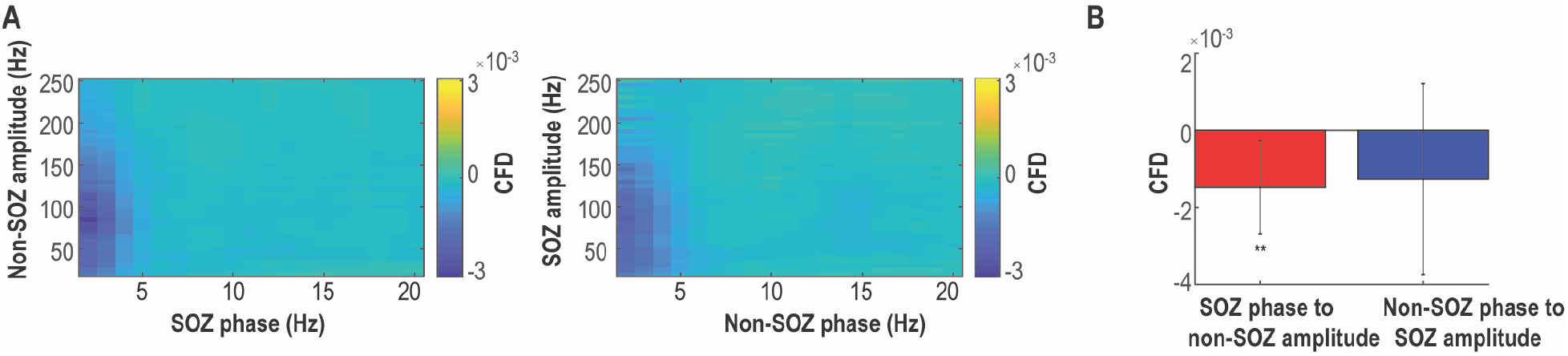
Cross-frequency information flow. (A) Grand averaged SOZ phase to non-SOZ amplitude CFD (left panel) and non-SOZ phase to SOZ amplitude CFD (right panel) across all electrode pairs and patients. (B) Grand averaged CFD after the k-means clustering procedure. The SOZ phase to non-SOZ amplitude CFD is significant compared to zero. The error bar represents standard deviation. ** *P*<0.01.

### Higher excitability in non-SOZ versus SOZ revealed by 1/f power slope

The differences in information flow between SOZ and non-SOZ could be related to alternation in excitation/inhibition balance. Based on computational modeling, it has been shown that E:I ratio could be estimated from the 1/f power slope, in which the more negative power slope is associated with less excitability (*51*). Therefore, we investigated the 1/f power slope as an indicator of E:I balance. After computing the power spectrum between 1 and 250 Hz at each electrode, the 1/f power slope was derived with the FOOOF algorithm (*52*). Among 184 SOZ and 899 non-SOZ electrodes, we found that power slopes of SOZ were significantly more negative than non-SOZ electrodes (two-sample *t*-test, *p*<10^−9^, Fig. 4A). On a single patient basis, 33 out of 43 patients had a more negative power slope in SOZ (Fig. 4B). Taken together, SOZ had a more negative 1/f power slope in comparison to non-SOZ, probably reflecting the higher excitability in non-SOZ.

**Fig. 4.**
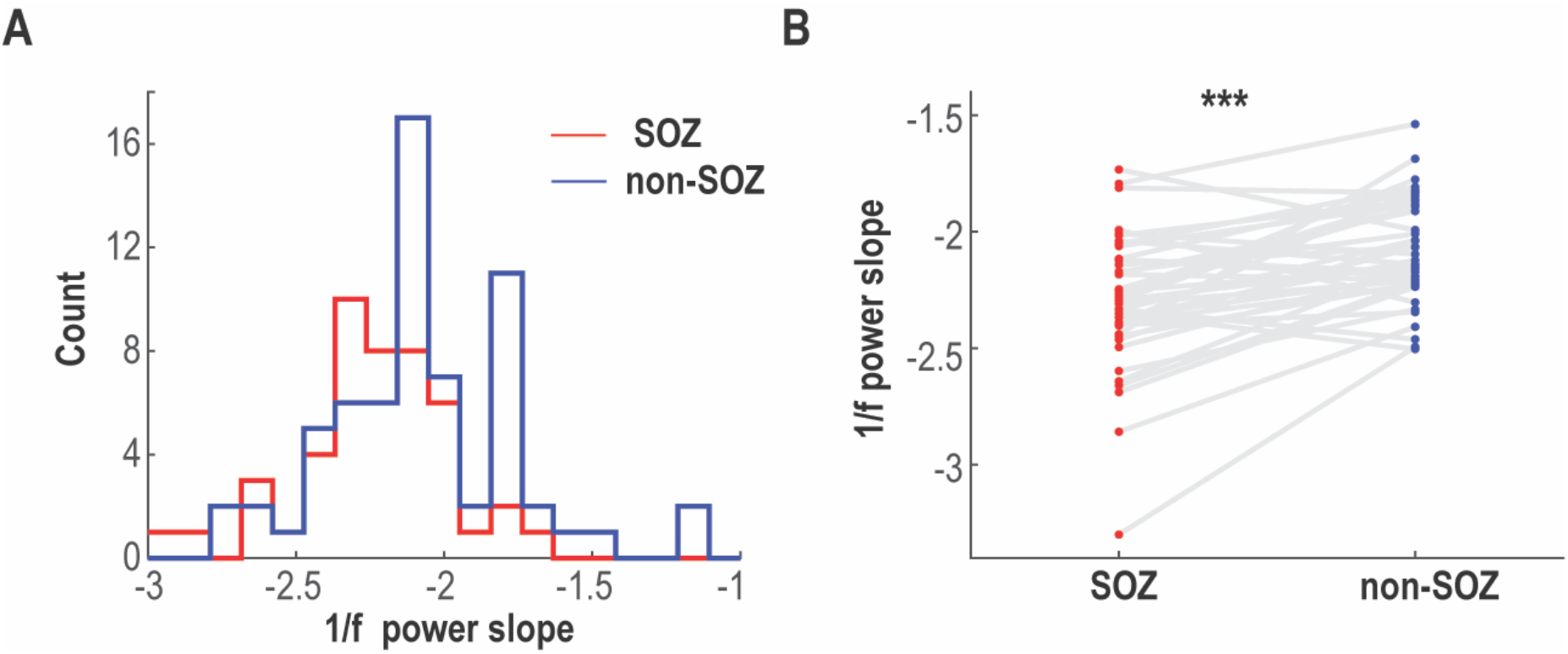
1/f power slope. (A) Distribution of 1/f power slope values shifts leftward (more negative) in SOZ (red) vs. non-SOZ (blue) electrodes. (B) Individual-patient comparison of averaged 1/f power slopes between SOZ (red) and non-SOZ (blue), each patient represented by a pair of connected dots showing the majority of patients (76.7%) had more negative slopes in SOZ compared to non-SOZ. *** *p*<0.001.

### Larger information flow asymmetry between SOZ and non-SOZ is associated with favorable seizure outcome

Next, we investigated the association between connectivity and seizure outcome. Of these 43 patients, there were 22 patients with Engel I outcome (51.2%), 8 patients with Engel II outcome (18.6%), 8 patients with Engel III outcome (18.6%), and 5 patients with Engel IV outcome (11.6%). We classified Engel I outcome as seizure-free and Engel II-IV outcome as the non-seizure free outcome. Notably, the majority of seizure free outcome patients had resection (14 patients) and ablation (5 patients) treatments. In the neural data, we found significant differences in within-frequency bidirectional information flow between SOZ and non-SOZ in the broadband frequency range in the seizure-free patients. At the same time, there was no significant difference in non-seizure free patients (Fig. 5A). After averaging over the broadband frequencies, the differences in seizure outcome were driven by weaker SOZ to non-SOZ information flow strength and stronger non-SOZ to SOZ information flow strength in seizure-free patients (Fig. 5B). Note that we did not find such significant differences in the cross-frequency information flow. Taken together, these suggested that larger within-frequency information flow asymmetry between SOZ and non-SOZ was associated with favorable seizure outcome.

**Fig. 5.**
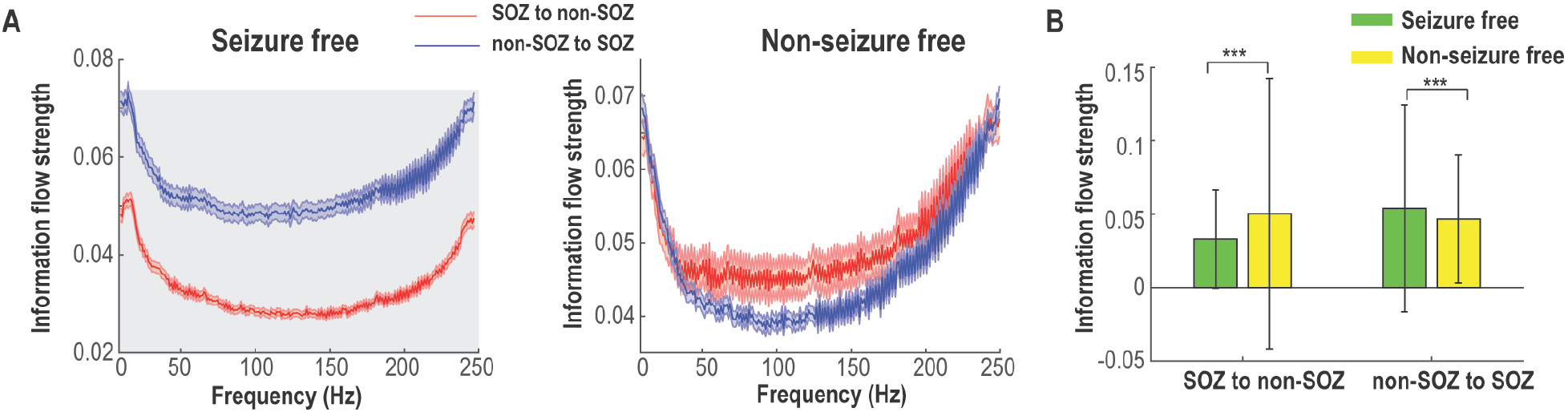
Association of information flow with post-seizure outcome. (A) Within-frequency information flow between SOZ and non-SOZ according to seizure outcome. The shaded gray area indicates significant difference at the *P*=0.05 level after FDR correction. Data are shown in mean and standard error. (B) Averaged bidirectional within-frequency information flow between SOZ and non-SOZ over the broadband frequencies in A. SOZ to non-SOZ information flow strength was significantly weaker, but non-SOZ to SOZ information flow strength was substantially stronger in seizure-free than non-seizure free patients. Error bar represents standard deviation. \*\*\**P*<0.001.

### Individual predictions of SOZ and seizure outcome with random forest classifier

Lastly, we utilized the random forest classifier to predict: 1) whether an individual electrode is likely to be SOZ; 2) whether the patient will be seizure-free. Based on the statistical results above, within-frequency interaction was significantly different in both information flow between SOZ and non-SOZ comparison (Fig. 2) and seizure-free outcome versus non-seizure free outcome comparison (Fig. 5). We only used the broadband within –frequency information flow as feature inputs into the random forest classifier to increase interpretability. More specifically, the strength of within-frequency inward information flow at each electrode was the feature for SOZ prediction, while the strength of mean non-SOZ to SOZ information flow for each patient was the feature for seizure outcome prediction. As shown in Fig. 6A, the model demonstrated an accuracy of 0.85 and an AUC of 0.94 in predicting SOZ (Precision: 0.91; Recall: 0.93) vs non-SOZ (Precision: 0.84; Recall: 0.78). For seizure outcome prediction, the model achieved an accuracy of 0.77 and an AUC of 0.78 when SOZ was identified by clinicians (Precision: [0.73 0.82]; Recall: [0.86 0.67] for seizure free and non-seizure free outcomes) (Fig. 6B). If SOZ was estimated by our prediction model, the seizure outcome prediction accuracy was 0.72 with an AUC of 0.75 (Precision: [0.71 0.77]; Recall: [0.74 0.67] for seizure free and non-seizure free outcomes) (Fig. 6C). Overall, these findings suggest that the combination of random forest and resting-state connectivity may help identify SOZ and predict seizure outcome at the individual level with satisfactory accuracy.

**Fig. 6.**
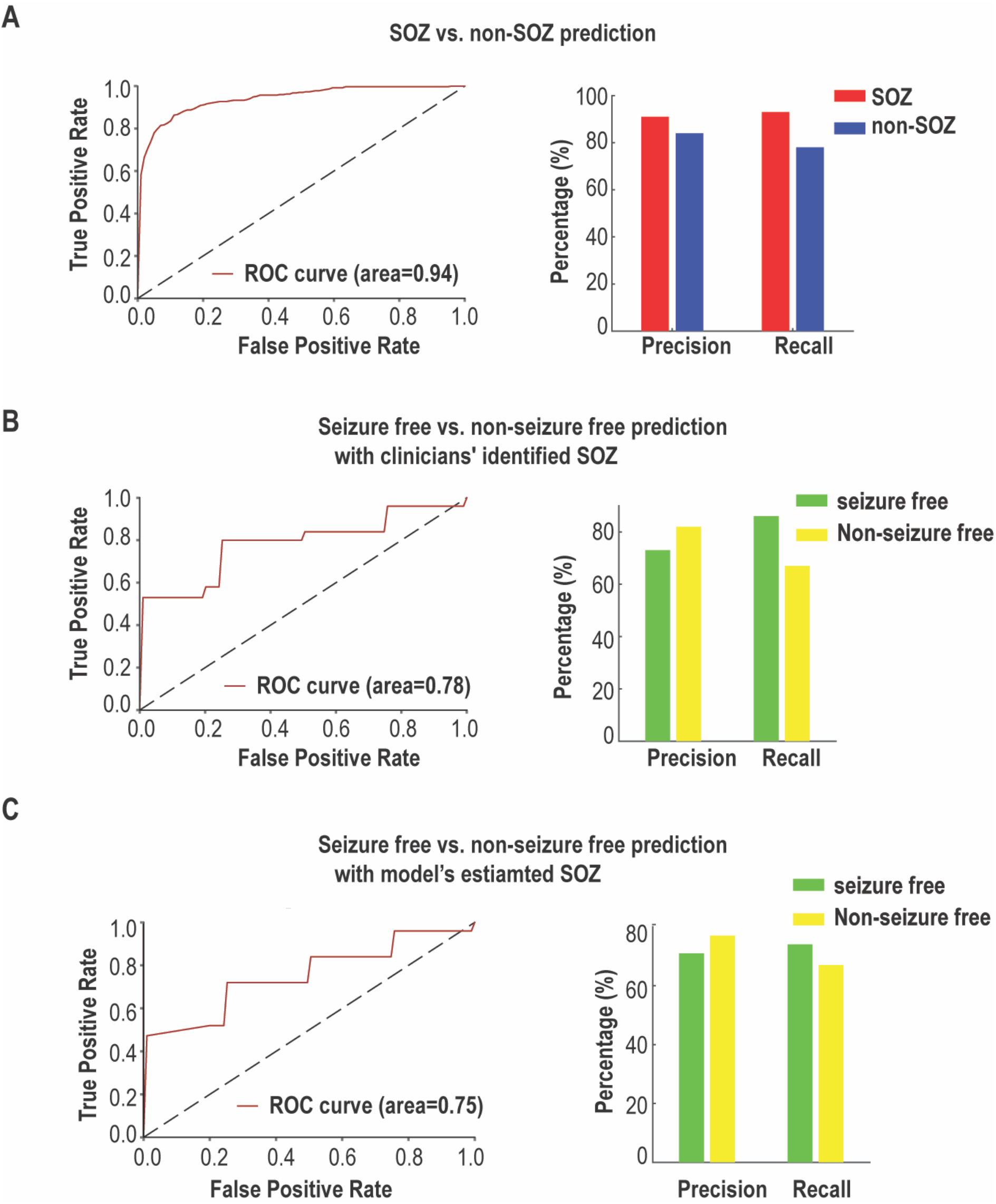
Performance of SOZ and seizure outcome predictions at the individual level. (A) SOZ versus non-SOZ prediction. Receiver-operating characteristic (ROC) curves show the true-positive and false-positive rates in predicting SOZ vs. non-SOZ. The area under the curve (AUC) is 0.94. Precision = True Positive / (True Positive + False Positive); Recall= True Positive / (True Positive + False Negative); (B) Similar to (A) but for prediction of seizure outcome with clinicians’ identified SOZ, i.e., seizure-free versus non-seizure free. (C) Similar to (B) but with model’s estimated SOZ, where only 10-min resting state SEEG data were used.

## Discussion

Overall, our results suggest that the dominant information flow is always from non-SOZ to SOZ at multiple frequencies in the interictal period, which is probably due to the higher excitability in non-SOZ regions. Moreover, larger information flow asymmetry between SOZ and non-SOZ is associated with favorable seizure outcome. By incorporating both resting-state connectivity and random forest classifier, it is possible to localize SOZ and predict seizure outcome at the individual level with satisfactory accuracy.

### Multiple oscillatory push-pull antagonisms underlying the epilepsy resting-state

We hypothesized that there is competition in information flow between SOZ and non-SOZ underlying epilepsy resting state (Fig. S2). There are two possibilities under our hypothesis: 1) SOZ tends to send more information to non-SOZ; 2) non-SOZ sends more information to SOZ. Our data support the latter since we found that the dominant multilayer neuronal interaction is always from non-SOZ to SOZ during the resting-state. More specifically, SOZ received more information flow from non-SOZ in a broadband frequency in the within-frequency network (Fig. 2), while the HFA of non-SOZ sent information to the LFA of SOZ in the cross-frequency network (Fig. 3). Interestingly, a previous study has shown that the focal seizure propagation dynamic was constrained by push-pull antagonisms between SOZ and non-SOZ (*38*). The modulation of non-SOZ primarily determines whether the seizure propagates or not. During the resting state, non-SOZ may send more directional information flow to SOZ to prevent seizure spread, probably reflecting the widespread network inhibition (*58*). Furthermore, more considerable asymmetry in within-frequency information flow between SOZ and non-SOZ (weaker SOZ to non-SOZ information flow and stronger non-SOZ to SOZ information flow) was associated with favorable seizure outcome. These might suggest that the less capacity for SOZ to spread, the more suppression from non-SOZ to SOZ, the better seizure outcome.

### Disrupted excitation-inhibition balance in epilepsy

Neural circuits rely on a dynamic E:I balance, and the balance of E:I interaction is critical for neuronal homeostasis and neural oscillation formation (*59, 60*). Emerging evidence indicates that E:I balance has dynamically fluctuated with neural computation, task demands, and cognitive states (*52*). More dramatic changes and aberrant E:I patterns are implicated in neurological disorders such as epilepsy (*61, 62*). The computation model developed by Gao et al. suggested that the E:I ratio can be quantified from the power spectrum, with a flatter 1/f power slope (more positive value) indicating a higher E:I ratio (*51*). This was supported by the evidence that 1/f power slope tracked the propofol-induced global inhibition, in which significant slope decrease was observed during anesthesia when compared to awake. Our data showed a more negative power slope in SOZ compared to non-SOZ (Fig. 4), probably reflecting low excitability in SOZ but high excitability in non-SOZ during the resting state. The disrupted excitation-inhibition balance might be linked to our findings in information flow. The non-SOZ with high excitability could be the source of information sender to SOZ with low excitability, explaining why the dominant information is from non-SOZ to SOZ. Moreover, a more negative power slope in SOZ was mainly driven by the high-frequency activity (30-250 Hz, Fig. S3), suggesting its frequency-specific effect.

### Long-range communication disruption of high-frequency activity in the epilepsy network

Synchronization between neuronal populations is critical for information transfer between brain areas (*63*). Theoretical and experimental evidence has shown that synchronization between neuronal oscillations depend on the axonal conduction delays, so LFA are generally more stably synchronized over long distance than HFA (*64, 65*). Surprisingly, our data challenged this classical view and demonstrated HFA exhibiting long-range communication both within SOZ, within non-SOZ (Fig. S4), and between SOZ and non-SOZ (Fig. 7). We estimated the DTF between all SEEG electrode pairs from 1 to 250 Hz and divided them into three groups based on distances. The DTFs were first averaged over all patients in three quartiles of inter-electrode distances. The mean DTFs increased from 1 to 6 Hz in all distance quartiles and then decayed to 80 Hz. However, throughout the 80-250 Hz HFA, inter-electrode DTFs started to increase again and exhibited a peak at around 240 Hz. Note that the long-range communication coordinated by HFA was shown in a recent study in healthy regions (*66*). Here, we extended the previous findings and provided the first evidence of long-range communication disruption of HFA between SOZ and non-SOZ in the pathological epilepsy network. The long-range neuronal communication of HFA could arise in large-scale network. This is probably because the joint roles of local synchronization and high collective firing rates enable local pyramidal cell populations with largely increased efficiency in regulating their post-synaptic targets in distant regions (*67, 68*). Moreover, the significant differences between SOZ and non-SOZ directional connectivity strength were found in short-range (<33 mm) and medium-range (between 33 mm and 60 mm) distance, but not in the long-range (between 60 mm and 159 mm) distance (Fig. 7), suggesting that disruption of HFA in the epilepsy network may limit to 60 mm away from SOZ.

**Fig. 7.**
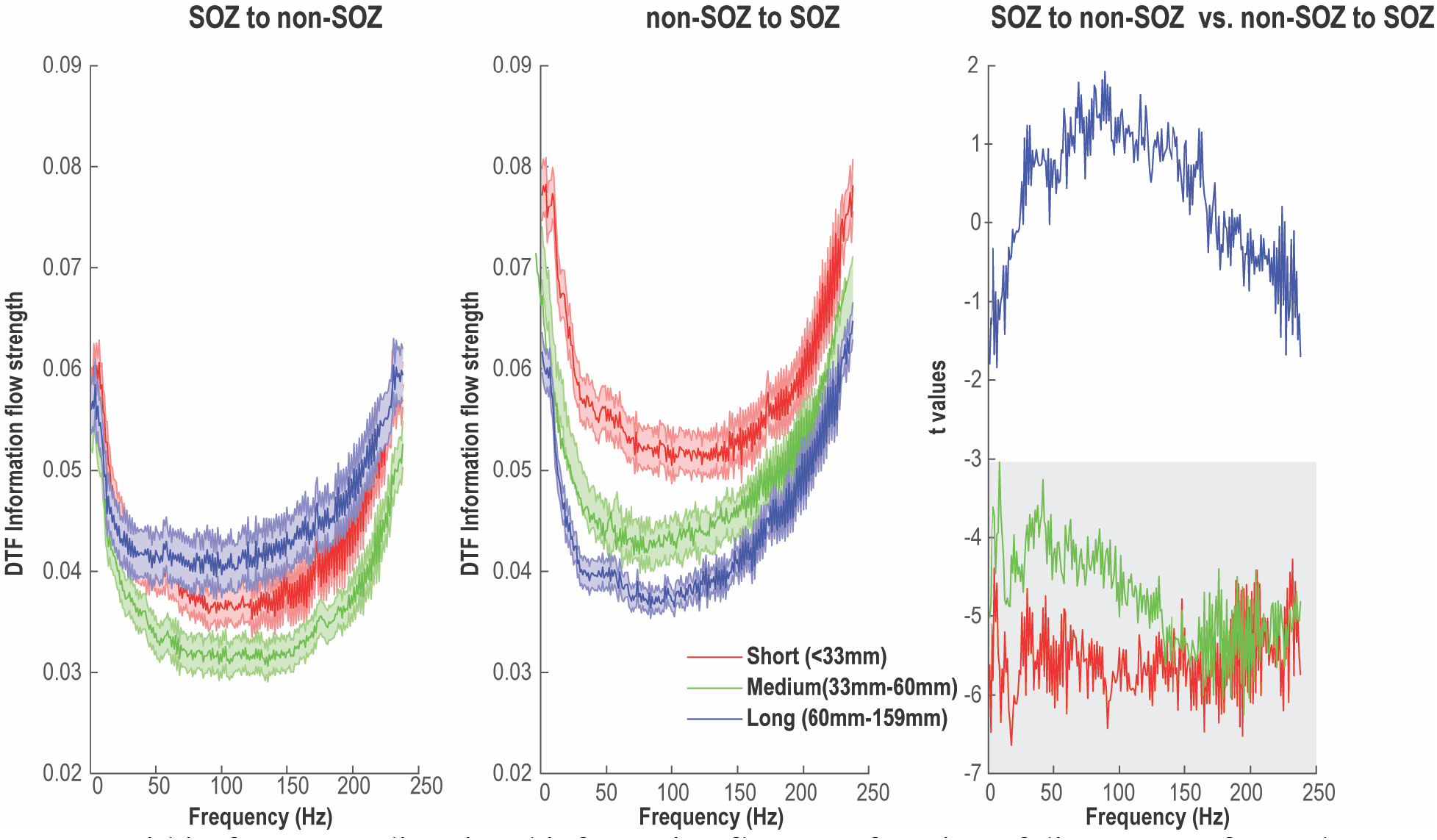
Within-frequency directional information flow as a function of distance. Left panel: Mean SOZ to non-SOZ DTF of all electrode pairs in distance-range quartiles. Middle panel: Mean non-SOZ to SOZ DTF of all electrode pairs in distance-range quartiles. Right panel: Statistical difference between SOZ to non-SOZ DTF and non-SOZ to SOZ DTF in distance-range quartiles. Significant area at *p*=0.05 level after FDR correction is marked in shadow. Data are shown in mean and standard error.

### Contributions of resting-state data in clinical decision-making

Ultimately, this study aims to improve seizure outcome of epilepsy patients, which largely relies on the precise delineation of epileptic networks. Here, we demonstrated that our approach could predict both SOZ and seizure outcome with about 80% accuracy in a large cohort of 43 drug resistant focal epilepsy patients using 10 minutes of interictal intracranial EEG resting-state data. The interpretations of ictal data have limitations, mainly imposed by accelerated meditation changes during the intracranial study and electrode coverage leading to sampling biases that may affect localization accuracy (*69, 70*). Moreover, it is challenging to capture all types of seizures during hospitalization (*71*). For example, one study showed that approximately one-third of bilateral temporal lobe epilepsy patients required more than four weeks of recordings to capture bilateral independent seizures (*72*). In addition, seizure clusters may provide discordant data that may misdirect interpretation and surgical treatment (*73*).

The underlying pathology of seizure generation most likely involves both abnormal brain structures and aberrant connections among these regions, leading to large-scale network disruptions (*31*). The aberrant network connection could be studied under resting-state, and many resting-state intracranial EEG studies have shown overwhelmingly enhanced connectivity involving EZ and surrounding regions (*74, 75*). More recently, two SEEG resting-state studies suggested the possibility to predict SOZ/EZ at the individual electrode level (*15, 16*). Goodale et al. computed 8-12 Hz alpha-band imaginary coherence across all electrodes using 2 min resting-state SEEG data in a cohort of 15 adult focal epilepsy patients (*15*). Six functional connectivity measures were incorporated in the logistic regression model to predict epileptogenicity of individual regions, and their model showed an AUC of 0.78 and an accuracy of 80.4%. Narasimhan et al. investigated 25 focal epilepsy patients with 2 minutes of resting-state, artifact-free SEEG data, and calculated three non-directed connectivity measures and four directed measures in the alpha band (*16*). Logistic regression was further applied to generate a predictive model of ictogenicity with an AUC of 0.88 and an accuracy of 84.3%. In our work, we used 10 minutes resting-state SEEG data to predict both SOZ and seizure outcome in a cohort of 43 epilepsy patients. We investigated both within-frequency and cross-frequency directional connectivity network during a wide frequency range from 1 Hz – 250 Hz. To tackle the problem of severely imbalanced data between SOZ and non-SOZ, a balanced random forest model was introduced by optimizing the cost function and the sampling technique. Our results, obtained from a relatively large patient population using a network connectivity approach, demonstrated enhanced performance of localizing SOZ with an AUC of 0.94 and an accuracy of 0.85. Besides, we examined the E:I ratio by computing the 1/f power slope and provided deeper mechanistic insights between the E:I alteration and aberrant connectivity in the resting-state epilepsy network. Furthermore, by utilizing directional connectivity network information, we made important advancement to predict seizure outcome with satisfactory accuracy. Prospective validation of our findings would pave the way to reducing traditional prolonged seizure recordings, leading to shorter hospitalizations and improved patient care.

### Study Limitations

One limitation of this study is that we used only the clinical SOZ to determine the EZ concept. This is a limitation because the SOZ can be a subset of the EZ, suggested but the fact that only a fraction of our patients achieved seizure-freedom (i.e., Engel I). However, our goal was to evaluate how the interictal resting state can accurately localize the SOZ identified from the intracranial ictal recordings, and this is a significant contributory factor. Another limitation stems from the short 10-min artifact and spike-free interictal data selection for our analysis, while studies have shown temporal variations of the epileptiform activity during longer periods of time (*76, 77*). We acknowledge that these epochs represent only snapshots of the 24-hour brain activity, and that further research is needed to investigate how behavior states (sleep/awake) influence our findings. However, our findings are stable and robust under different durations with 5 min, 15 min and 30 min (Fig. S5). Finally, we only used within-frequency information flow as features to make an individual prediction of seizure outcome for simplicity and more straightforward interpretation. It is important to note that a few clinical variables such as absence of generalized tonic-clonic seizures and presence of hippocampal atrophy were significantly associated with seizure remission (*78*). Future studies will be needed to develop multivariate outcome prediction models by taking clinical variables into account.

## Conclusions

We have investigated the aberrant information flow and the disrupted excitation-inhibition balance in epilepsy resting state. We found that the higher excitability in non-SOZ versus SOZ regions could be linked to the dominant information flows from non-SOZ to SOZ at multiple frequencies, probably reflecting insufficient excitability to initiate seizure and widespread network inhibition to prevent seizure initiation. Moreover, stronger information flow from non-SOZ to SOZ was found in seizure free outcome patients compared to non-seizure free outcome patients. In combination with the balanced random forest machine learning model and resting-state connectivity, localization of SOZ and seizure outcome prediction without long-term recordings may supplement traditional interpretation of SEEG and help identify epilepsy treatment targets, thus improving patient care and treatment outcome.

## Data Availability

All data produced in the present work is contained in the manuscript.

## Acknowledgment

This work was supported in part by NIH grants EB021027, NS096761, MH114233, EB029354, AT009263 (B.H.). We are grateful to Dr. Abbas Sohrabpour and Zhengxiang Cai for helpful discussions on data analysis. We thank participating patients and their families whose involvement and sacrifice made this work possible. We also acknowledge selfless dedication and invaluable efforts of the University of Pittsburgh Comprehensive Epilepsy Center (UPCEC) team and particularly the staff of the UPMC Presbyterian University Hospital (PUH) Epilepsy Monitoring Unit (EMU).

## Author Contributions

H.J. and B.H. conceived the idea. H.J. and B.H. designed the study and wrote the manuscript draft. H.J. analyzed the data. M.R., A.B., V.K., A.U. collected the data. S. Y., and V.K. prepared and pre-processed the data. M.R., A.B. and V.K. revised the manuscript. All the authors discussed the results and contributed to the manuscript. B.H. supervised the research.

## References

1. J. W. Sander, S. D. Shorvon, Epidemiology of the epilepsies. J. Neurol. Neurosurg. Psychiatry 61, 433–443 (1996).

2. D. J. Englot, E. F. Chang, Rates and predictors of seizure freedom in resective epilepsy surgery: an update. Neurosurg. Rev. 37, 389–404; discussion 404-385 (2014).

3. H. O. Luders, I. Najm, D. Nair, P. Widdess-Walsh, W. Bingman, The epileptogenic zone: general principles. Epileptic Disord. 8 Suppl 2, S1–9 (2006).

4. F. Rosenow, H. Lüders, Presurgical evaluation of epilepsy. Brain 124, 1683–1700 (2001).

5. R. M. Richardson, Decision Making in Epilepsy Surgery. Neurosurg. Clin. N. Am. 31, 471–479 (2020).

6. P. Chauvel, J. Gonzalez-Martinez, J. Bulacio, Presurgical intracranial investigations in epilepsy surgery. Handb. Clin. Neurol. 161, 45–71 (2019).

7. L. Jehi, The Epileptogenic Zone: Concept and Definition. Epilepsy Curr. 18, 12–16 (2018).

8. M. Cossu, G. Lo Russo, S. Francione, R. Mai, L. Nobili, I. Sartori, L. Tassi, A. Citterio, N. Colombo, M. Bramerio, C. Galli, L. Castana, F. Cardinale, Epilepsy surgery in children: results and predictors of outcome on seizures. Epilepsia 49, 65–72 (2008).

9. J. P. Mullin, M. Shriver, S. Alomar, I. Najm, J. Bulacio, P. Chauvel, J. Gonzalez-Martinez, Is SEEG safe? A systematic review and meta-analysis of stereo-electroencephalography-related complications. Epilepsia 57, 386–401 (2016).

10. D. J. Englot, A modern epilepsy surgery treatment algorithm: Incorporating traditional and emerging technologies. Epilepsy Behav. 80, 68–74 (2018).

11. J. P. Mullin, M. Shriver, S. Alomar, I. Najm, J. Bulacio, P. Chauvel, J. Gonzalez-Martinez, Is SEEG safe? A systematic review and meta-analysis of stereo-electroencephalography–related complications. Epilepsia 57, 386–401 (2016).

12. B. C. Jobst, F. Bartolomei, B. Diehl, B. Frauscher, P. Kahane, L. Minotti, A. Sharan, N. Tardy, G. Worrell, J. Gotman, Intracranial EEG in the 21st Century. Epilepsy Curr. 20, 180–188 (2020).

13. J. Bancaud, R. Angelergues, C. Bernouilli, A. Bonis, M. Bordas-Ferrer, M. Bresson, P. Buser, L. Covello, P. Morel, G. Szikla, A. Takeda, J. Talairach, Functional stereotaxic exploration (SEEG) of epilepsy. Electroencephalogr. Clin. Neurophysiol. 28, 85–86 (1970).

14. S. Rheims, P. Ryvlin, Patients’ safety in the epilepsy monitoring unit: time for revising practices. Curr. Opin. Neurol. 27, 213–218 (2014).

15. S. E. Goodale, H. F. J. Gonzalez, G. W. Johnson, K. Gupta, W. J. Rodriguez, R. Shults, B. P. Rogers, J. D. Rolston, B. M. Dawant, V. L. Morgan, D. J. Englot, Resting-State SEEG May Help Localize Epileptogenic Brain Regions. Neurosurgery 86, 792–801 (2020).

16. S. Narasimhan, K. B. Kundassery, K. Gupta, G. W. Johnson, K. E. Wills, S. E. Goodale, K. Haas, J. D. Rolston, R. P. Naftel, V. L. Morgan, B. M. Dawant, H. F. J. Gonzalez, D. J. Englot, Seizure-onset regions demonstrate high inward directed connectivity during resting-state: An SEEG study in focal epilepsy. Epilepsia 61, 2534–2544 (2020).

17. S. Lagarde, N. Roehri, I. Lambert, A. Trebuchon, A. McGonigal, R. Carron, D. Scavarda, M. Milh, F. Pizzo, B. Colombet, B. Giusiano, S. Medina Villalon, M. Guye, C. G. Benar, F. Bartolomei, Interictal stereotactic-EEG functional connectivity in refractory focal epilepsies. Brain 141, 2966–2980 (2018).

18. G. Buzsáki, A. Draguhn, Neuronal Oscillations in Cortical Networks. Science 304, 1926–1929 (2004).

19. H. Jiang, T. Popov, P. Jylanki, K. Bi, Z. Yao, Q. Lu, O. Jensen, M. A. van Gerven, Predictability of depression severity based on posterior alpha oscillations. Clin. Neurophysiol. 127, 2108–2114 (2016).

20. C. Wilke, G. Worrell, B. He, Graph analysis of epileptogenic networks in human partial epilepsy. Epilepsia 52, 84–93 (2011).

21. M. Guirgis, Y. Chinvarun, M. del Campo, P. L. Carlen, B. L. Bardakjian, Defining regions of interest using cross-frequency coupling in extratemporal lobe epilepsy patients. Journal of Neural Engineering 12, (2015).

22. S. P. Burns, S. Santaniello, R. B. Yaffe, C. C. Jouny, N. E. Crone, G. K. Bergey, W. S. Anderson, S. V. Sarma, Network dynamics of the brain and influence of the epileptic seizure onset zone. Proc. Natl. Acad. Sci. U. S. A. 111, E5321–E5330 (2014).

23. A. N. Khambhati, K. A. Davis, T. H. Lucas, B. Litt, D. S. Bassett, Virtual Cortical Resection Reveals Push-Pull Network Control Preceding Seizure Evolution. Neuron 91, 1170–1182 (2016).

24. G. Worrell, J. Gotman, High-frequency oscillations and other electrophysiological biomarkers of epilepsy: clinical studies. Biomark. Med. 5, 557–566 (2011).

25. K. J. Staley, F. E. Dudek, Interictal spikes and epileptogenesis. Epilepsy Curr. 6, 199–202 (2006).

26. M. Amiri, B. Frauscher, J. Gotman, Phase-Amplitude Coupling Is Elevated in Deep Sleep and in the Onset Zone of Focal Epileptic Seizures. Front. Hum. Neurosci. 10, (2016).

27. N. Zaher, A. Urban, A. Antony, C. Plummer, A. Bagic, R. M. Richardson, V. Kokkinos, Ictal Onset Signatures Predict Favorable Outcomes of Laser Thermal Ablation for Mesial Temporal Lobe Epilepsy. Front. Neurol. 11, 595454 (2020).

28. O. Grinenko, J. Li, J. C. Mosher, I. Z. Wang, J. C. Bulacio, J. Gonzalez-Martinez, D. Nair, I. Najm, R. M. Leahy, P. Chauvel, A fingerprint of the epileptogenic zone in human epilepsies. Brain 141, 117–131 (2018).

29. A. Bernasconi, Connectome-based models of the epileptogenic network: a step towards epileptomics? Brain 140, 2525–2527 (2017).

30. P. Jiruska, M. de Curtis, J. G. Jefferys, C. A. Schevon, S. J. Schiff, K. Schindler, Synchronization and desynchronization in epilepsy: controversies and hypotheses. J. Physiol. 591, 787–797 (2013).

31. J. Engel, Jr., P. M. Thompson, J. M. Stern, R. J. Staba, A. Bragin, I. Mody, Connectomics and epilepsy. Curr. Opin. Neurol. 26, 186–194 (2013).

32. S. B. Tomlinson, B. E. Porter, E. D. Marsh, Interictal network synchrony and local heterogeneity predict epilepsy surgery outcome among pediatric patients. Epilepsia 58, 402–411 (2017).

33. J. Engel, Jr., P. Van Ness, T. Rasmussen, L. Ojemann, Outcome with respect to epileptic seizures. In Engel J Jr, ed. Surgical Treatment of the Epilepsies, 609–621 (1993).

34. S. Lagarde, S. Buzori, A. Trebuchon, R. Carron, D. Scavarda, M. Milh, A. McGonigal, F. Bartolomei, The repertoire of seizure onset patterns in human focal epilepsies: Determinants and prognostic values. Epilepsia 60, 85–95 (2019).

35. S. Singh, S. Sandy, S. Wiebe, Ictal onset on intracranial EEG: Do we know it when we see it? State of the evidence. Epilepsia 56, 1629–1638 (2015).

36. B. He, L. Astolfi, P. A. Valdes-Sosa, D. Marinazzo, S. Palva, C. G. Benar, C. M. Michel, T. Koenig, Electrophysiological Brain Connectivity: Theory and Implementation. IEEE Trans. Biomed. Eng., (2019).

37. M. Kaminski, M. Ding, W. A. Truccolo, S. L. Bressler, Evaluating causal relations in neural systems: granger causality, directed transfer function and statistical assessment of significance. Biol. Cybern. 85, 145–157 (2001).

38. H. Jiang, Z. Cai, G. A. Worrell, B. He, Multiple Oscillatory Push-Pull Antagonisms Constrain Seizure Propagation. Ann. Neurol. 86, 683–694 (2019).

39. H. T. Jiang, A. Bahramisharif, M. A. J. van Gerven, O. Jensen, Measuring directionality between neuronal oscillations of different frequencies. NeuroImage 118, 359–367 (2015).

40. M. J. Kaminski, K. J. Blinowska, A New Method of the Description of the Information-Flow in the Brain Structures. Biol. Cybern. 65, 203–210 (1991).

41. M. Kaminski, M. Z. Ding, W. A. Truccolo, S. L. Bressler, Evaluating causal relations in neural systems: Granger causality, directed transfer function and statistical assessment of significance. Biol. Cybern. 85, 145–157 (2001).

42. C. Wilke, W. van Drongelen, M. Kohrman, B. He, Neocortical seizure foci localization by means of a directed transfer function method. Epilepsia 51, 564–572 (2010).

43. C. Wilke, W. van Drongelen, M. Kohrman, B. He, Identification of epileptogenic foci from causal analysis of ECoG interictal spike activity. Clin. Neurophysiol. 120, 1449–1456 (2009).

44. L. Ding, G. A. Worrell, T. D. Lagerlund, B. He, Ictal source analysis: localization and imaging of causal interactions in humans. NeuroImage 34, 575–586 (2007).

45. R. Oostenveld, P. Fries, E. Maris, J. M. Schoffelen, FieldTrip: Open source software for advanced analysis of MEG, EEG, and invasive electrophysiological data. Comput. Intell. Neurosci. 2011, 156869 (2011).

46. H. Park, D. S. Lee, E. Kang, H. Kang, J. Hahm, J. S. Kim, C. K. Chung, H. Jiang, J. Gross, O. Jensen, Formation of visual memories controlled by gamma power phase-locked to alpha oscillations. Sci. Rep. 6, 28092 (2016).

47. R. F. Helfrich, M. Huang, G. Wilson, R. T. Knight, Prefrontal cortex modulates posterior alpha oscillations during top-down guided visual perception. Proc. Natl. Acad. Sci. U. S. A. 114, 9457–9462 (2017).

48. R. F. Helfrich, B. A. Mander, W. J. Jagust, R. T. Knight, M. P. Walker, Old Brains Come Uncoupled in Sleep: Slow Wave-Spindle Synchrony, Brain Atrophy, and Forgetting. Neuron 97, 221–230 e224 (2018).

49. J. Zheng, K. L. Anderson, S. L. Leal, A. Shestyuk, G. Gulsen, L. Mnatsakanyan, S. Vadera, F. P. Hsu, M. A. Yassa, R. T. Knight, J. J. Lin, Amygdala-hippocampal dynamics during salient information processing. Nat Commun 8, 14413 (2017).

50. G. Nolte, A. Ziehe, V. V. Nikulin, A. Schlogl, N. Kramer, T. Brismar, K. R. Muller, Robustly estimating the flow direction of information in complex physical systems. Phys. Rev. Lett. 100, 234101 (2008).

51. R. Gao, E. J. Peterson, B. Voytek, Inferring synaptic excitation/inhibition balance from field potentials. NeuroImage 158, 70–78 (2017).

52. T. Donoghue, M. Haller, E. J. Peterson, P. Varma, P. Sebastian, R. Gao, T. Noto, A. H. Lara, J. D. Wallis, R. T. Knight, A. Shestyuk, B. Voytek, Parameterizing neural power spectra into periodic and aperiodic components. Nat. Neurosci. 23, 1655–U1288 (2020).

53. L. Breiman, Random Forests. Machine Learning 45, 5–32 (2001).

54. C. Chao, L. Andy, B. Leo, Using Random Forest to Learn Imbalanced Data. University of California, Berkeley, 1–12 (2004).

55. N. V. Chawla, K. W. Bowyer, L. O. Hall, W. P. Kegelmeyer, SMOTE: Synthetic minority over-sampling technique. J Artif Intell Res 16, 321–357 (2002).

56. G. Lemaitre, F. Nogueira, C. K. Aridas, Imbalanced-learn: A Python Toolbox to Tackle the Curse of Imbalanced Datasets in Machine Learning. J Mach Learn Res 18, (2017).

57. L. A. Baccala, K. Sameshima, Partial directed coherence: a new concept in neural structure determination. Biol. Cybern. 84, 463–474 (2001).

58. D. J. Englot, H. Blumenfeld, Consciousness and epilepsy: why are complex-partial seizures complex? Prog. Brain Res. 177, 147–170 (2009).

59. B. V. Atallah, M. Scanziani, Instantaneous Modulation of Gamma Oscillation Frequency by Balancing Excitation with Inhibition. Neuron 62, 566–577 (2009).

60. J. Touboul, A. Destexhe, Can Power-Law Scaling and Neuronal Avalanches Arise from Stochastic Dynamics? PLoS One 5, e8982 (2010).

61. C. Symonds, Excitation and inhibitiion in epilepsy. Brain 82, 133–146 (1959).

62. L. R. Gonzalez-Ramirez, O. J. Ahmed, S. S. Cash, C. E. Wayne, M. A. Kramer, A biologically constrained, mathematical model of cortical wave propagation preceding seizure termination. PLoS Comput. Biol. 11, e1004065 (2015).

63. M. Bonnefond, S. Kastner, O. Jensen, Communication between Brain Areas Based on Nested Oscillations. eNeuro 4, (2017).

64. G. Arnulfo, J. Hirvonen, L. Nobili, S. Palva, J. M. Palva, Phase and amplitude correlations in resting-state activity in human stereotactical EEG recordings. NeuroImage 112, 114–127 (2015).

65. D. A. Leopold, Y. Murayama, N. K. Logothetis, sVery slow activity fluctuations in monkey visual cortex: Implications for functional brain imaging. Cereb. Cortex 13, 422–433 (2003).

66. G. Arnulfo, S. H. Wang, V. Myrov, B. Toselli, J. Hirvonen, M. M. Fato, L. Nobili, F. Cardinale, A. Rubino, A. Zhigalov, S. Palva, J. M. Palva, Long-range phase synchronization of high-frequency oscillations in human cortex. Nat Commun 11, 5363 (2020).

67. W. Singer, Neuronal Synchrony: A Versatile Code for the Definition of Relations? Neuron 24, 49–65 (1999).

68. M. B. Khamechian, V. Kozyrev, S. Treue, M. Esghaei, M. R. Daliri, Routing information flow by separate neural synchrony frequencies allows for “functionally labeled lines” in higher primate cortex. Proceedings of the National Academy of Sciences 116, 12506–12515 (2019).

69. S. Rheims, P. Ryvlin, Patients’ safety in the epilepsy monitoring unit: time for revising practices. Curr. Opin. Neurol. 27, 213–218 (2014).

70. R. Di Giacomo, R. Uribe-San-Martin, R. Mai, S. Francione, L. Nobili, I. Sartori, F. Gozzo, V. Pelliccia, M. Onofrj, G. Lo Russo, M. de Curtis, L. Tassi, Stereo-EEG ictal/interictal patterns and underlying pathologies. Seizure 72, 54–60 (2019).

71. P. A. Karthick, H. Tanaka, H. M. Khoo, J. Gotman, Could we have missed out the seizure onset: A study based on intracranial EEG. Clin. Neurophysiol. 131, 114–126 (2020).

72. D. King-Stephens, E. Mirro, P. B. Weber, K. D. Laxer, P. C. Van Ness, V. Salanova, D. C. Spencer, C. N. Heck, A. Goldman, B. Jobst, D. C. Shields, G. K. Bergey, S. Eisenschenk, G. A. Worrell, M. A. Rossi, R. E. Gross, A. J. Cole, M. R. Sperling, D. R. Nair, R. P. Gwinn, Y. D. Park, P. A. Rutecki, N. B. Fountain, R. E. Wharen, L. J. Hirsch, I. O. Miller, G. L. Barkley, J. C. Edwards, E. B. Geller, M. J. Berg, T. L. Sadler, F. T. Sun, M. J. Morrell, Lateralization of mesial temporal lobe epilepsy with chronic ambulatory electrocorticography. Epilepsia 56, 959–967 (2015).

73. M. A. Astrada, A. S. Marti, R. McLachlan, D. Diosy, S. Mirsattari, D. Steven, J. Burneo, Can We Accurately Lateralize the Epileptogenic Zone in Patients Who Have Seizure Clusters? A Study Using Stereoelectroencephalography (SEEG) (1710). Neurology 94, 1710 (2020).

74. G. Bettus, F. Wendling, M. Guye, L. Valton, J. Regis, P. Chauvel, F. Bartolomei, Enhanced EEG functional connectivity in mesial temporal lobe epilepsy. Epilepsy Res. 81, 58–68 (2008).

75. M. Holmes, B. S. Folley, H. H. Sonmezturk, J. C. Gore, H. Kang, B. Abou-Khalil, V. L. Morgan, Resting state functional connectivity of the hippocampus associated with neurocognitive function in left temporal lobe epilepsy. Hum. Brain Mapp. 35, 735–744 (2014).

76. S. V. Gliske, Z. T. Irwin, C. Chestek, G. L. Hegeman, B. Brinkmann, O. Sagher, H. J. L. Garton, G. A. Worrell, W. C. Stacey, Variability in the location of high frequency oscillations during prolonged intracranial EEG recordings. Nat Commun 9, 2155 (2018).

77. E. C. Conrad, S. B. Tomlinson, J. N. Wong, K. F. Oechsel, R. T. Shinohara, B. Litt, K. A. Davis, E. D. Marsh, Spatial distribution of interictal spikes fluctuates over time and localizes seizure onset. Brain 143, 554–569 (2019).

78. S. S. Spencer, A. T. Berg, B. G. Vickrey, M. R. Sperling, C. W. Bazil, S. Shinnar, J. T. Langfitt, T. S. Walczak, S. V. Pacia, S. Multicenter Study of Epilepsy, Predicting long-term seizure outcome after resective epilepsy surgery: the multicenter study. Neurology 65, 912–918 (2005).

